# Perceptions about male circumcision among HIV vaccine efficacy trial participants in Soweto, South Africa: a qualitative study

**DOI:** 10.1101/2023.05.03.23289466

**Authors:** Mbalenhle Sibiya, Fatima Laher, Mamakiri Mulaudzi, Lerato M. Makhale, Taibat Salami, Stefanie Hornschuh, Hong-Van Tieu, Janan J. Dietrich

## Abstract

Male circumcision has both health benefits and significance to some cultures. We sought to understand perceptions about male circumcision as part of the HIV prevention toolkit among participants enrolled in a preventive HIV vaccine efficacy trial in South Africa. We conducted a qualitative study with 28 people aged 18-35 years old who self-reported that they were not living with HIV, provided informed consent, and who were participating in the HVTN 702 vaccine efficacy trial in Soweto. Using a semi-structured guide, we facilitated four focus group discussions (FGDs) stratified by age, gender, and sexual orientation. FGDs were audio-recorded, transcribed verbatim, translated into English, and thematically analysed. Four main themes emerged. (1) Everyone had accurate knowledge about what male circumcision is, and some participants stated that it partially reduces acquisition of HIV and sexually transmitted infections. (2) There was an emerging distrust of cultural circumcision because of perceived lack of transparency and adverse events. (3) There was a perception that circumcision boosted masculinity. (4) The choice to circumcise was influenced by parents, family, and female partners. In conclusion, the study found that young South African HIV vaccine trial participants accurately understand the HIV prevention benefits of male circumcision, but uptake decisions are embedded within a context that is informed by culture, sexuality and masculinity norms and values.

## Introduction

Male circumcision, which is the removal of the penile foreskin, has been shown in clinical trials to reduce the risk of heterosexual acquisition of human immunodeficiency virus (HIV) among circumcised cisgender men,[1,2,3,4] as well as reducing the incidence of urinary tract infections, penile cancer, human papilloma virus (HPV), herpes simplex virus type 2 (HSV-2), and genital ulcer disease.[5] Although there are no data from randomized clinical trials, several observational studies have shown that male circumcision also partially protects against HIV acquisition for gay and bisexual males who predominantly or exclusively engage in insertive anal intercourse.[1,6]

Furthermore, there is evidence that male circumcision is associated with better health outcomes for female partners, including risk reduction of HPV, cervical dysplasia, cervical cancer, HSV-2, chlamydia, and syphilis, and there is limited data that male circumcision lowers HIV acquisition risk for female partners.[7]

In South Africa, male circumcisions are performed in multiple contexts. In this paper, we use the term ‘cultural circumcision’ to refer to traditional male circumcision performed within the cultural context of initiation schools,[8] and the term ‘voluntary medical male circumcision’ to refer to circumcision in the medical context by surgical methods or non-surgical devices.[9] Cultural male circumcisions, which are not regulated by the government and are generally performed in non-medical settings, have been criticised for high complication rates including dehydration, infection, disfigurement and death,[10] and interventions to improve their safety have had limited success.[11] In Lesotho, where most male circumcisions are performed in the traditional context, a study found that circumcisions performed in initiation schools do not have the same medical benefits as those performed in the surgical context, despite no evidence of behavioural disinhibition, and this is likely because the entire foreskin is not removed.[8]

Beyond health benefits, male circumcision holds significant cultural significance in South Africa. The cultural practice of male circumcision is likely to have been more pervasive historically.[12] Currently, its practice is maintained amongst some cultures, including Sotho and Xhosa men who regard it as a sacred religious practice, a preparation for acquired manhood, and a rite of passage into adulthood.[13] Performed in initiation schools, traditional ritual circumcision establishes a link between male participants of an age cohort.[12] Although initiates are required to maintain secretiveness about what is taught in initiation schools, one author suggested a recent idea has emerged among the Xhosa tradition that initiation gives men the unrestricted right to sex rather than introducing concepts about sexual responsibility.[14]

Male medical circumcision is an important biomedical tool for HIV prevention. In our study, we explored the attitudes toward male circumcision as an HIV prevention tool, including its cultural context. Here we use the term “male” to refer to those persons assigned male sex at birth.

## Methods

### Setting and participants

While the preventive HVTN 702 HIV vaccine trial was ongoing, we conducted a qualitative study at one of the trial sites, the Perinatal HIV Research Unit (PHRU).[15] The site is in Soweto, South Africa, a township with an estimated population size of 1,271,628 people.[16]

The eligibility criteria for our qualitative study were participants enrolled in HVTN 702 (which, in brief, required participants to be healthy, 18-35 years old, at risk for HIV acquisition, not living with HIV, and willing to discuss HIV risk reduction), who stated they were currently not living with HIV and provided written voluntary informed consent.

### Procedures

Using purposive sampling, potential participants were recruited in 2018 at the trial site. Recruitment methods included posting paper flyers on PHRU noticeboards, and the qualitative study staff approaching people during their study visits at the site. Interested participants were pre-screened for age and enrolment status in HVTN 702 using a questionnaire on an electronic computer tablet. Staff then telephoned potentially eligible participants to schedule a focus group discussion (FGD). There was a limit of 10 participants per FGD to preserve the potential for inter-participant communication and airing of a range of views.

On the day of the FGD, after obtaining written consent, participants completed a demographic questionnaire in English on paper. Multilingual study staff read the questionnaire, explaining in English and/or local languages (i.e., English, Zulu, and Sotho) as preferred. The demographic questionnaire assessed date of birth, gender identity, sexual orientation, marital status, highest level of education completed, HIV risk perception of self and of partners, and with whom the participant lived.

Finally, they participated in the FGD. Trained facilitators conducted four FGDs stratified by age, gender, and sexual orientation: (i) heterosexual and bisexual women aged 25-35 years; (ii & iii) heterosexual men aged 25-35 years; and (iv) men aged 18-35 years who have sex with men (MSM). The facilitators used a semi-structured guide containing open-ended questions about male circumcision with probes designed to elicit discussion (Table 1).

**Table 1.** Summary of semi-structured FGD guide questions and probes

| **Area** | **Main questions** | **Probes** |
| --- | --- | --- |
| Knowledge on and attitudes toward voluntary male medical circumcision | What do you know about voluntary male medical circumcision? | What did you learn about voluntary male medical circumcision from your participation in the vaccine trial? |
| Perceptions on culture and voluntary male medical circumcision | Do you think culture influences voluntary male circumcision? |  |
| Perceptions on voluntary male medical circumcision and HIV prevention | What is your understanding on the role of voluntary male medical circumcision in HIV prevention? | Do you think that voluntary male medical circumcision protects one from HIV infection? |
| Role of gender in choices about voluntary male medical circumcision | For female participants with male sexual partners: How easy or difficult do you think it is for females to talk about voluntary male medical circumcision with their male partners? | ● Have any of your sexual partners been circumcised?<br>● How important do you think it is for your partner to get circumcised?<br>● How likely are you to refer your partner/s for voluntary male medical circumcision? |
|  |  | Any reasons why your partner may not undergo voluntary male medical circumcision? |

FGDs lasted up to 110 minutes and were held at PHRU in a private room separate from the HVTN 702 clinic. A local multilingual female facilitator and an English-speaking female co-facilitator led FGDs with women. A local multilingual male facilitator and a multilingual female co-facilitator led FGDs with men. Facilitators were experienced in qualitative research and trained in HIV prevention. FGDs were conducted in a mix of local languages audio-recorded, transcribed verbatim, translated into English, and validated against the audio recording.

Participants were reimbursed ZAR150 (∼USD12) for transport costs.

In HVTN 702, trained study staff collected the circumcision status of all enrolled male participants by examination, and classified their answers as no circumcision, full circumcision (full removal of the foreskin covering the penile glans), and partial circumcision (partial removal of the foreskin). Staff entered the data into a case report form on an electronic data capture system.

### Ethical and community considerations

The University of the Witwatersrand Human Research Ethics Committee approved the study. The HVTN 702 protocol team and the PHRU HIV prevention community advisory board reviewed this qualitative study and provided input.

### Data analysis

Demographic data for participants in the qualitative study were entered into an online database on SurveyPlanet.[17] Circumcision data collected in HVTN 702 for all male participants enrolled at the Soweto site were copied from the baseline circumcision examination case report form into the iMedidata electronic database used for this study. Descriptive statistics and frequencies were calculated using Microsoft Excel.

The FGDs were transcribed verbatim by trained staff. Qualitative data were analysed using thematic analysis.[18] Two analysts read the transcripts to become familiar with the data. The primary analyst conducted a line-by-line analysis assigning codes and categories to text. Codes and categories were not pre-determined, but rather emerged from the data. Both analysts refined the codes, categories and themes through multiple discussions and re-readings of the transcripts, until they agreed on a final list. Quotations were selected to exemplify the themes; the anonymity of the speaker was preserved by crediting them to their focus group (e.g. F25-35 means the focus group of females aged 25 to 35 years old) and not to individuals.

## Results

### Participant demographics

Of 81 HVTN 702 participants approached, 28 (35%) participated in one of the four focus groups. Nine participants were heterosexual and bisexual women aged 25-35 years, 14 were heterosexual men aged 25-35 years who were divided equally into two groups, and 5 were men and a transgender female aged 18-35 years who have sex with men. The median age was 28 years. Nine (32%) identified as women, 18 (64%) as men, and 1 (4%) as transgender female. The sexual orientation of 5 (18%) participants was homosexual, 1 (4%) was bisexual, and 22 (79%) were heterosexual. Twenty-six (93%) participants were single, 1 (4%) was widowed, and 1 (4%) was married. Three (11%) participants had not completed a high school education. For perception of HIV acquisition risk, 19 (68%) reported high/moderate risk, and 14 (50%) reported that their partners were at high/moderate risk. Four (14%) participants lived alone, and the remainder lived with family.

### Circumcision status in HVTN 702

In HVTN 702, 106 males were enrolled at the Soweto site, of which 53 (50%) were not circumcised at baseline. The proportions of no circumcision, full circumcision, and partial circumcision were similar between heterosexual and non-heterosexual identifying males as shown in Table 2.

**Table 2.**
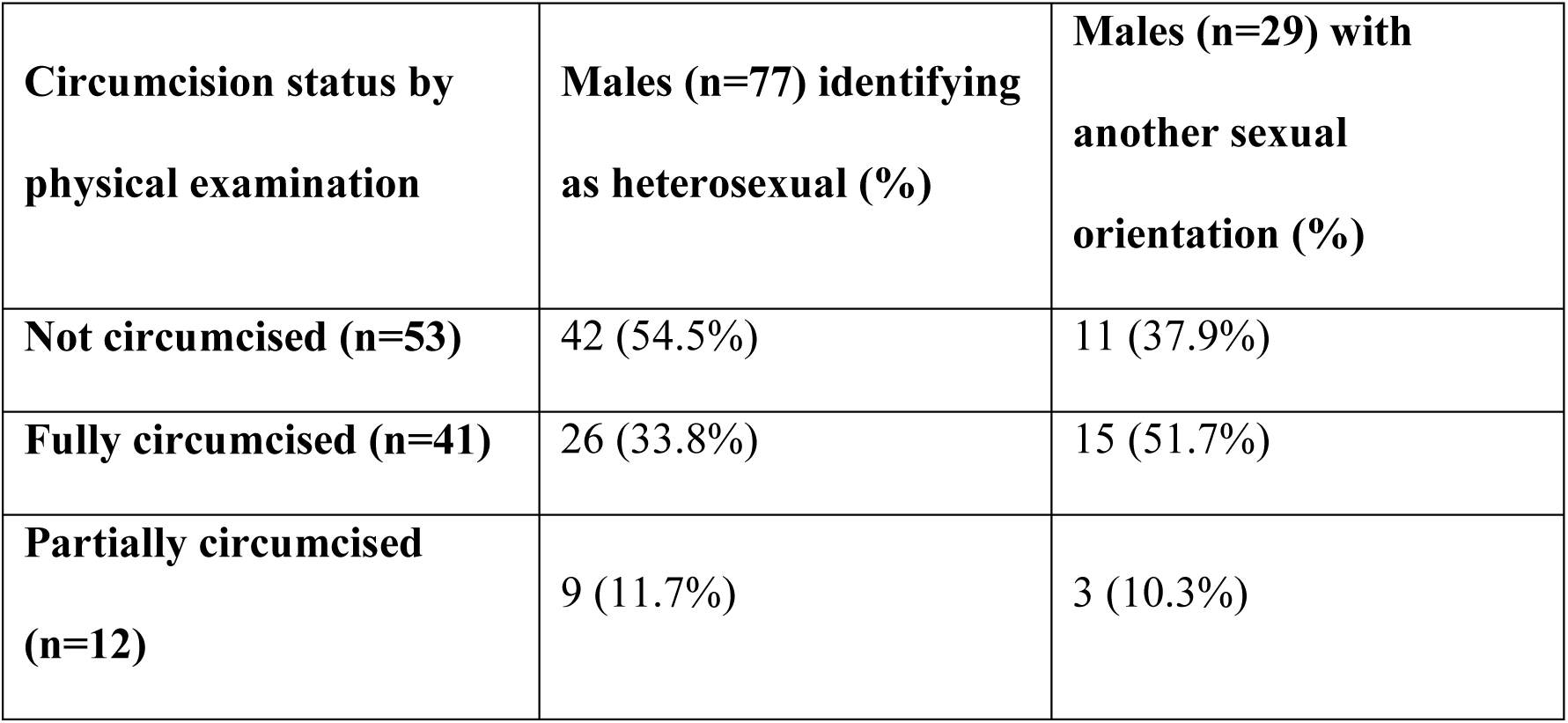
Circumcision status of 106 males enrolled in HVTN 702 at the Soweto trial site.

### Qualitative results

Four main themes emerged from the FGDs.

#### (i) Knowledge about male circumcision and reduced acquisition of HIV and sexually transmitted infections (STIs)

Most participants agreed that the removal of penile foreskin contributed to reduced chances of HIV and STI acquisition by circumcised males. Most participants presented knowledge that male circumcision does not eliminate the risk of acquiring HIV entirely.

*“But then again also male circumcision helps, it doesn’t prevent, it doesn’t like 100% prevent you from getting HIV… But it minimizes the risk.” (F25-35)*

*“I know that if you want to reduce the risks of you getting STIs you can get circumcised.” (M25-35)*

*“I know that MMC [Medical Male Circumcision], right, it reduces the chance of one being infected…” (MSM 18-35)*

MSM participants volunteered that the importance of circumcision extended to men in same sex relationships to reduce HIV acquisition, but a few perceived that receptive partners would not have to get circumcised unless they preferred to do so. Most MSM participants thought that insertive partners would gain protection by circumcision, but receptive partners would not.

*“I think when it comes to like gay men, it is a matter of preference, but you need to do it, but if you are a bottom [receptive partner], like it is not a must-do, because you do not use it (penis)…” (MSM 18-25)*

Heterosexual and MSM male participants perceived that the foreskin encourages infections. Women and MSM participants perceived that circumcised men were more hygienic. Some female participants perceived that being uncircumcised was a barrier for males to use condoms.

*“There’s nothing more frustrating as, [laughing] when you must roll the penis – shift it [foreskin] that way and the condom the other way. And that thing plus a condom is what hurts actually.” (F 25-35)*

*“…some boys don’t know how to wash properly; you find that the foreskin is closed off…” (F25-35)*

#### (ii) Emerging concerns about cultural circumcision

All participants said there were differences between cultural and medical circumcisions. It was said that cultures differ with regard to circumcision requirements. For example, they noted that Xhosa and Sesotho cultures required circumcision, but there were also some people within those cultures who did not regard circumcision as a cultural requirement. Heterosexual and MSM Male participants said that the option of how to get circumcision was determined by the region where they come from.

*“…since like I grew up in Johannesburg, so I do not think my parents will exactly have a problem with me being circumcised, but culturally, going back to the rural, they would not approve of me being circumcised…” (MSM18-35)*

Although some participants preferred cultural circumcision, most volunteered reasons why they felt uncertain or doubtful about cultural circumcision. Women had concerns that the tools used for cultural circumcision were not correct for the task or were not sterilised. Participants across all FGDs mentioned that they perceived there was insufficient information about what happens during cultural circumcision and that the secrecy around cultural circumcision made them prefer medical circumcision. Some heterosexual males perceived it as limiting that the clinic provides only health education, and not cultural information about masculinity. Male participants agreed that culture forbade males to disclose details about the experience of African cultural circumcision, but both male and female participants shared that the teachings at initiation schools included ideas of masculinity. Although MSM and female participants dismissed the teachings as gender-stereotyped, most heterosexual males valued the teachings as providing important guidance for life.

First participant: “Because now going to the mountains, what is it that they are using… or they use an axe or they use a…”

Second participant: “Razor”.

First participant: “If perhaps it was used by another person when you think about it, I think on this one they use whatever is available. Do you understand and those things are not hygienic and they not surgically cleaned.” *(F25-35)*

“The only thing I’m saying is that there is a difference between and man who went to the clinic and a man who went to the initiation school. In the initiation school there are things that you are taught about, manhood stuff in case you have a son or daughter in future, how you should raise them and how you should handle them and then when you go to the clinic it’s based on your health that this is what you are supposed to do from 3 to 6 weeks until you heal. After you heal then you have to find out information for yourself”. *(M25-35)*

*“If you ask a person when they come back and you ask them to tell you what happened there [at initiation school], then, they told me not to say anything.” (F25-35)*

*“The thing is there is something, there is something that I cannot disclose. I for one I went to the mountain, so I cannot disclose everything and put it down here in the open.” (M25-35)*

Both heterosexual males and MSM participants stated that they preferred medical circumcision at the local clinics because of the transparency of information, monitoring during healing, and provision of analgesia. Most heterosexual males perceived value in the endurance of pain and ‘initiation school teachings’ said to be part of the African cultural male circumcision experience, but said that it would be acceptable if medical clinics did not anaesthetise. Participants were concerned that people are injured and die during cultural circumcision.

*“I prefer clinical procedure… rather than the initiation l school, because of their case is going to the initiation school whereby people are contracting HIV/AIDS, they cut the using the same blade…there are stories and when you go to the clinic, the first thing they do they test you for HIV AIDS before providing counselling, after counselling they cut you with clean blades. Everything is perfect at the clinic and they monitor you after the procedure, you come back after 3 days to check how you are healing.”(M25-35)*

*“… I have information as a clinic, information at the clinic after they circumcise you, you are going to drink pills, you are going to stay inside your house for six weeks.” (M25-35)*

*“That is why I suggested that it is best both ways to limit the death. What causes people to die in the mountains (cultural circumcision) is they do not listen to what they are told and mostly again it’s dehydration, because you are not allowed to drink water. I will stop there; I will not go further [to describe the ritual].’’ (M25-35)*

Participants also stated a concern that the same blade is used on everyone during cultural circumcision, a practice they regarded as non-hygienic and linked with the possibility of HIV transmission. They stated that this was different from local medical clinics, which they preferred, because a new blade was used for each person and HIV testing was offered.

Some suggested that cultural and medical circumcision practices be combined to limit the number of deaths and HIV acquisition. They envisioned a service where people would first get medically circumcised at the local clinic, then go to initiation schools to learn about their culture and masculinity. Simultaneously, there was a concern that it would not be culturally acceptable to combine practices.

*“…then I would suggest that both be exercised, maybe you could start at a clinic and then later on go to the mountain, because what they teach you there at the mountain is not exactly the same as what they teach you at the clinic, there is other things that a lot of us we went to the mountain, have information about woman, sex, how to live, how to survive and how to take things as a man.” (M25-35)*

*“…Because remember it is not going to the mountains or to initiation, it is not necessarily just about getting the laws of tradition, but to also go through the pain of you know, your penis being cut, you know with an okapi [pocket knife], you know you getting that pain… Yes, so, so if you are saying you will go to the mountain after you have been to the clinic I do not, I do not for see that happening, that is just my honest opinion.” (M25-35)*

#### (iii) Circumcision for boosting masculinity

The discussions revealed a variety of reasons why men choose to circumcise. There was a perception among men of all sexual orientation that the benefits of medical male circumcision are rarely what motivates most men to circumcise. The decision to circumcise was mostly tied to boosting an image of masculinity. This idea of masculinity was discussed more in the context of cultural circumcision: men were expected to take part in it to honour family expectations so that they could gain social status and respect. Men of all sexual orientations said it was linked with the idea of manliness that was restricted to heterosexuality. A male participant perceived that girls were brought to initiation schools after cultural circumcision to test masculinity.

*“…and in some places in the initiation school, because you are discharged to go home…they bring girls for you to test how your penis works and that your parents did not waste their money.” (M25-35)*

Women were doubtful that cultural circumcision teaches boys to be men. Some participants said that the main motivation to choose cultural circumcision is to transition from being a boy to being a man. Some participants across all FGDs perceived that those who did not undergo cultural circumcision would never be recognized within their culture as a man. It was said that in some places, circumcised men are perceived by society to be more “manly’’ than uncircumcised men, which motivates the choice to circumcise.

*“…. what they teach you there at the mountain (cultural circumcision) is not exactly the same as what they teach you at the clinic. There’s other things that a lot of us, we went to the mountain (cultural circumcision), have information about: women, sex, how to live, how to survive and how to take things as a man.*” *(M25-35)*

Male participants perceived that cultural circumcision increased penile size and libido.

*“Seriously, since I’m back from the mountain (Cultural circumcision) and have endured the cold, I am a beast in bed.’’ (M25-35)*

Participants perceived that medical circumcision makes the penis smoother, cleaner and larger, and men said that it prevents weak erections, all perceived as benefits. Most female participants mentioned that they enjoyed having sex with men who were medically circumcised because the penis was smoother and cleaner.

#### (iv) External influences in circumcision choice

Across all groups, it emerged that the decision to circumcise is not always made by the individual. Influence may come from external factors like culture, parents and female sexual partners. Participants mentioned that in some locales in rural areas, circumcision was practised as a rite of passage and parents made it mandatory to go to initiation schools. In contrast, those from urban areas were free to choose whether they wanted to be circumcised or not.

*“Yes, I hear you, and I support him, as he says maybe about 10% does traditional circumcision but according to how I was brought up including my grandfather and great grandfather, I went for the traditional circumcision and so I also wish for my child to also go for the traditional circumcision. He will collect all the health information from the clinic after he has been to the mountain (for cultural circumcision).’*’ *(M25-35)*

Both heterosexual and MSM participants mentioned that they would get circumcised in order to please their parents and family.

“*It varies according to like different beliefs, like Xhosa people as opposed to Sotho people, they are more strict when it comes to, like, going to the mountain (cultural circumcision) and doing it, because they feel like it is right of passage, do you understand? So even when you are a gay person, you also have to think of your family and you have to honour your family, I think.” (MSM18-35)*

*“…but then if I am forced to do it [cultural circumcision], under cultural influence, then yes, I have to undergo that…” (MSM18-35)*

Female participants mentioned that women influenced their male partners and family members to get circumcised. A female participant stated that she took all the male family members to the clinic to get medically circumcised. Women stated a preference for having sex with circumcised partners, and one female participant mentioned how she took her partner to the local clinic to get medically circumcised to improve his sexual performance:

*“I took him by the hand and said, you know what, if you gonna continue [claps hands] having sex with me, let’s go… Get circumcised then you will be alright…” (F25-35)*

## Discussion

Our qualitative study showed that, although there was consensus that male circumcision was a partially effective method to prevent HIV acquisition and STIs, more importance was placed on its perceived cultural role in establishing heteronormative masculinity and gaining societal status and respect. Males of all sexual orientations perceived that there are safety benefits of medical compared to cultural circumcision.

We found that participants had accurate knowledge about the benefits of male circumcision to men’s health, including prevention of HIV acquisition and STIs. Although we did not ask specifically about benefits to female partners, no participant volunteered any knowledge about it. It is likely that female benefits of male circumcision constitute a knowledge gap, which may be valuable to address with women to influence demand creation. MSM participants showed nuanced understanding that circumcision could partially protect insertive but not receptive partners from HIV acquisition. Generally, participants were not aware of the implications for protection of partial versus full circumcision.

A study conducted in the Western Cape province of South Africa found that there was less accurate knowledge about male circumcision.[19] Another study conducted in the KwaZulu Natal province of South Africa[20] found that Zulu men perceived circumcision as unnecessary and had some misconceptions about the mechanism of protection offered by circumcision. However, our findings support those from Uganda[21] that found accurate knowledge on the topic, but also cautioned that accurate knowledge about medical circumcision was not necessarily related to uptake. In Zimbabwe, knowledge about medical circumcision and HIV acquisition increased with age, education level, and media exposure.[22]

Most African research on this topic has focused on understanding the perspectives of heterosexual individuals. Our study adds to the literature by also documenting the perceptions of African MSM. HIV incidence is markedly higher among MSM compared to heterosexual men,[23] and therefore it is relevant to understand the knowledge of efficacious prevention methods among MSM. In our study, we found that although MSM participants demonstrated knowledge of the HIV prevention effect of male circumcision, there was a misperception that circumcision should follow sex role aggregation: only males who mainly practice penetrative anal sex should be circumcised. A similar finding was reported in China,[24] where circumcision uptake is low amongst MSM,[25] and males who mainly practice receptive anal sex perceive themselves as having low risk for HIV acquisition and regard circumcision as unnecessary.

Cultural circumcision is perceived with varying levels of trust. In this study, participants expressed some concerns about cultural circumcision, including morbidity and mortality risks, HIV transmission, and mandates of secrecy. Some of these medical concerns are upheld by data, which have shown increased complication rates in cultural compared to medical circumcisions, including hospitalisation, penile damage, and penile amputation.[26] In other research conducted in eastern and southern Africa, men reported abusive treatment as a concern.[27]

In contrast, in the Eastern Cape province of South Africa, there was a high level of trust in cultural circumcision, where participants attributed deaths not to the cultural practitioner, but to the interference from people external to the cultural practice.[28] Other authors noted community concerns that cultural circumcision introduced initiates to incorrect behaviours, such as encouraging substance use.[27] Our findings show that people would send their children for cultural circumcision if they had gone but it also supports results from other studies that some older men were reluctant to send their boys to the initiation school because they did not trust that these schools could teach their children how to become respectful men. Some of these men reported that young males who come from initiation schools showed new undesirable behaviours such as substance and drug use. Other studies have reported a decline in the practice of cultural circumcision, with parents preferring medical male circumcision for their children.[29] In our study, some participants did not perceive cultural circumcision as necessary within the urban community. The mandate of secrecy makes it hard for people within the community who have never had the experience of cultural circumcision to perceive it as safe.

We found that boosting masculinity was a major motivating factor for circumcision uptake. This phenomenon has been reported previously as culture-specific: for example, it has been reported that in the Xhosa culture in South Africa, circumcision affords men greater social status.[30,31] In addition to respect, the idea of masculinity also extended to sexuality: women in this study voiced a favourable perception of having sex with circumcised men.[32,33] Men reported that circumcision improved their sexual performance, which has been noted as an important demand creation message in other studies with similar outcomes.[34]

Multiple factors are known to influence the decision to circumcise males medically or culturally in Africa: family members, female partners,[35] cultural norms, and peer pressure.[36,37] In this study, participants reported that MSM participated in cultural circumcision because their families pressured them. Another South African study reported that parents would send their MSM children for cultural circumcision with the hope that it would convert them to heterosexuality, and the MSM would participate in cultural circumcision in hopes of being accepted by their communities and being given the social status and privileges given to heterosexual males who are culturally circumcised but not to boys or women.[38]

A limitation of our study is that the knowledge of medical circumcision displayed by the participants in this study is likely to be, in part, a reflection of routine risk reduction counselling offered in the vaccine trial. Therefore, this study may overestimate knowledge when compared to general populations who would not typically receive frequent counselling. A second limitation is that we did not collect the circumcision status of only the men involved in this study, and we do not have longitudinal circumcision data to understand how people’s uptake and perceptions changed over time. Although FGDs are subject to social desirability bias, a strength of our study design is that we grouped demographically similar participants and were able to compare findings across the different cohorts.

## Conclusions

Although the medical industry views male circumcision primarily as an intervention associated with HIV, STI, and other medically related prevention benefits, we showed that young South African men participating in HIV vaccines trials instead embed their uptake decisions within a context that is informed by culture, sexuality, and masculinity norms and values.

## Data Availability

The qualitative data illustrating the findings of the study are presented as participant quotes within the paper. The raw interview transcripts contain information that could potentially compromise participant privacy and will not be made public to ensure protection of participants.

## Acknowledgements

Author Contribution

MS analysed the data and wrote the draft. FL conceived the idea, contributed to study design, data analysis, and wrote the draft. MM led the protocol submission for local ethical committee approval, assisted with data analysis, and edited the manuscript. LMM collected the data, validated the results and edited the manuscript. TS wrote the grant application, collected the data, validated the results, and contributed to manuscript writing. SH managed the project and contributed to manuscript writing. HVT contributed to study design and manuscript editing. JJD designed the study, supervised the project, analysed the data and contributed to manuscript writing.

We thank the HIV Vaccine Trials Network (HVTN), Statistical and Data Management Center (SDMC), and the HVTN 702 Protocol Team for their support of this project. We are grateful to the study participants. We thank Putuke Kekana for assistance with conducting the FGDs. We thank Gail Broder and Michele P Andrasik for helpful comments on the manuscript.

## Disclaimer

The content is solely the responsibility of the authors and does not necessarily represent the official views of the NIAID, the National Institutes of Health (NIH), the Bill & Melinda Gates Foundation, or the South African Medical Research Council. GlaxoSmithKline Biologicals SA, maker of the vaccine tested in HVTN 702, was provided the opportunity to review a preliminary version of this manuscript, but the authors are solely responsible for final content and interpretation.

